# GEOMETRY OF THE CAROTID ARTERY AT BASELINE IMPROVES PREDICTION OF STENOSIS SEVERITY AT FOLLOW-UP

**DOI:** 10.64898/2025.11.28.25341216

**Authors:** Alexey Kamenskiy, William Poulson, Jason MacTaggart

**Author notes:** Correspondence and Reprints requests to: Alexey Kamenskiy, Ph.D., Department of Biomechanics, 6160 University of Nebraska Omaha, Omaha, NE 68182.

## Abstract

**Introduction:** Carotid artery geometry may contribute to the development of atherosclerotic carotid artery disease (ACD). Prior reports associated ACD with narrow bifurcation angles, tortuous Common Carotid Arteries (CCA), and large bulbs. Our objective was to determine whether deviations in these geometric features at baseline can help predict stenosis severity at follow-up.

**Methods:** A total of 20 human subjects (59±16 years old, range 31-85 years) with a 4.7±2.3 year (range 2.0-11.4 years) follow-up CTA imaging examination were identified, and n=36 carotid arteries that had no prior interventions were selected for analysis. Three-dimensional reconstructions of carotid arteries at baseline and follow-up were used to measure bifurcation angle, CCA tortuosity, bulb area, and stenosis severity, and these measurements were compared to previously reported values for healthy carotid arteries undergoing normal aging. Geometric factors were used along with clinical risk factors to predict severity of stenosis at follow-up using multivariate regression.

**Results:** Stenosis averaged 39±26% at baseline, and 44±28% at follow-up. The presence of coronary artery disease, hypertension, female gender, and deviation of bifurcation angle and tortuosity at baseline explained 67% of stenosis severity at follow-up. Clinical risk factors positively contributed to stenosis severity but when used alone explained only 47% of variability.

**Conclusions:** Narrow bifurcation angles and tortuous CCAs are associated with increased stenosis severity over time. Combining morphometric data with clinical risk factors may improve prediction of stenosis severity at follow-up, helping identify patients at higher risk for ACD and potentially allowing more accurate and personalized treatment recommendations.

## INTRODUCTION

Atherosclerotic carotid artery disease (ACD) is one of the leading causes of stroke, resulting in significant morbidity and mortality^1^. ACD is commonly associated with systemic risk factors, such as hypertension, smoking, hyperlipidemia, and diabetes mellitus, but the focal nature of the disease preferentially affecting the carotid bifurcation, suggests that geometric factors may also play a significant role. Blood flow disturbances and low shear stress in the carotid bulb that result from focal diameter increase in the carotid bifurcation are thought to contribute to formation of the atherosclerotic plaque^2^. However, all carotid arteries have bulbs, yet not all develop ACD. Furthermore, patients that develop unilateral disease are not uncommon, suggesting that local geometric features of the carotid artery bifurcation may contribute to initiation and progression of ACD^3^.

Prior study^3^ utilized healthy subjects and patients with unilateral ACD to demonstrate differences in normal and pathologic carotid artery geometries. Carotid arteries without ACD increased bifurcation angle, bulb diameter, and tortuosity with age, while presence of disease was associated with straight internal carotid arteries (ICA), and narrow bifurcation angles. While intriguing, these results were obtained from retrospective rather than longitudinal analysis of Computerized Tomography Angiography (CTA) scans, which limited their ability to predict disease progression.

The goal of the current study was to develop a method of predicting ACD progression by performing longitudinal assessment of carotid artery geometry using baseline and follow-up CTAs of patients in a wide range of ages and ACD severities to determine whether deviation in carotid geometric features from those of healthy carotid anatomies at baseline can help predict stenosis severity at follow-up.

## METHODS

### Patient population

Our entire institutional image database was searched for contrast-enhanced thin-section CT scans of the neck that were obtained in the same subjects at least two years apart. A total of 20 human subjects (59±16 years old, range 31-85 years, 7 male, 13 female, 18 Caucasian, 2 African American) with a 4.7±2.3 years (range 2.0-11.4 years) follow-up CTA imaging examination were identified, and n=36 carotid arteries that had no prior interventions were selected for analysis. Four carotid arteries were excluded because two were occluded at the time of the initial scan, and the other two had previous interventions. Average Body Mass Index (BMI) was 30±7 (range 20-47), 14 subjects had hypertension (HTN), 3 diabetes mellitus (DM), 14 had dyslipidemia, 3 had Coronary Artery Disease (CAD), and 14 used tobacco (average 16±9 pack/year history). CTA indications included neurologic symptoms (63%), carotid artery disease (33%), trauma (3%), and neck pain (1%). All patients with initial imaging obtained for neurological symptoms had a negative head CT scan demonstrating that these patients were asymptomatic for the carotid artery disease. All scans were retrospective, and the Institutional Review Board of the University of Nebraska Medical Center approved this study.

### Geometric features

Most CTAs were obtained with a 64-channel scanner Brilliance 64 (Philips Medical Systems, Cleveland, OH) with slice thicknesses of 0.625 mm and a resolution of 512×512 pixels (pixel size 0.391 mm). Bilateral carotid arteries on both baseline and follow-up scans were reconstructed from the aortic arch to the skull base using Mimics software (Materialize Co, Leuven, Belgium). Segmentations (Figure 1) were obtained using a combination of thresholding, region growing, and multi-slice edit techniques^3,4^. To reduce variability among CT scans, all segmentations were performed by a single operator.

**Figure 1.**
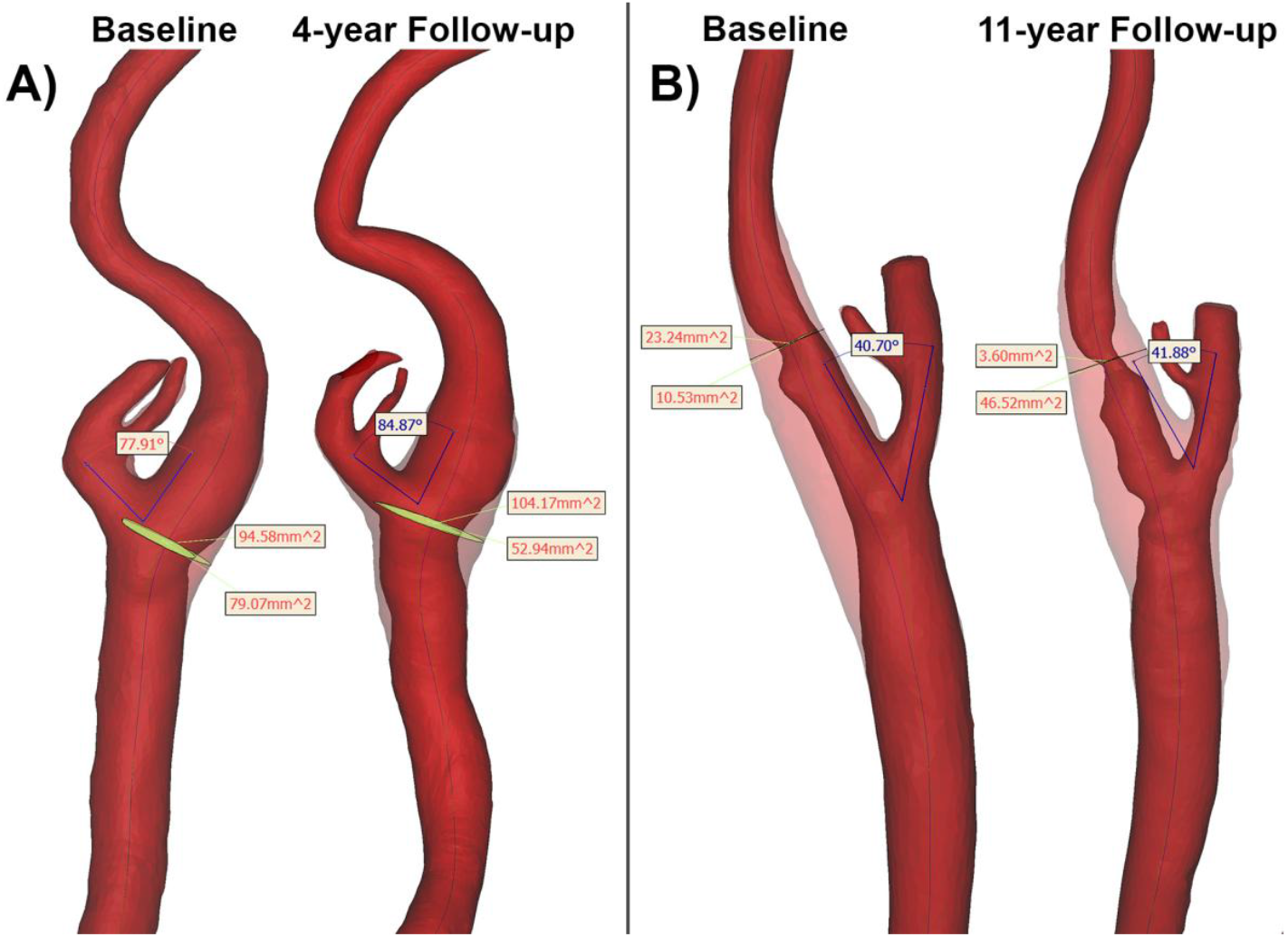
Baseline and follow-up carotid artery geometries demonstrating increased stenosis severity over 4-year (A) and 11-year (B) periods. Cross-sectional areas measured perpendicular to the outer wall and lumen centerlines were used to assess stenosis severity. In (A) stenosis severity increased from 16% to 49%, and in (B) from 55% to 92%. Angle of the carotid bifurcation was measured in 3D using the walls of the internal and external carotid arteries.

After completing 3D reconstructions, centerlines were built for both the arterial wall and lumen masks, and a number of geometrical features that were previously demonstrated important for ACD were measured^3^. These features included carotid artery bifurcation angle, common carotid artery (CCA) tortuosity, largest outer wall cross-sectional area of the carotid bulb and stenosis severity.

Angle of the carotid bifurcation was measured in 3D using the outer wall of the ICA and external carotid arteries (ECA) as demonstrated in Figure 1. This method was preferred over the centerline assessment because centerline position depends on bulb size, i.e. large bulbs lead to lateral deviation of the arterial centerlines resulting in the association of the bifurcation angle and stenosis severity^3^. CCA tortuosity was calculated as T_CCA_ = 1 − *l*/*L* where *l* is the shortest distance between the two locations, and *L* is the distance over the vessel centerline between the same points. For the left CCA, T_CCA_ was measured from the aortic arch to the point where CCA divides into the ICA and ECA, while for the right CCA T_CCA_ was measured from the brachiocephalic artery.

The largest cross-sectional area of the carotid bulb was measured in 3D perpendicular to the arterial centerline (Figure 1) instead of using 2D axial sections to account for the oblique nature of carotid bifurcations that often do not align with the CTA axial direction. In addition, we have used outer wall geometry to measure cross-sectional area of the bulb to avoid underestimation of bulb size due to stenosis. To avoid overestimation of cross-sectional area at the bifurcation of CCA into ECA and ICA, ECA was virtually removed along the line connecting the medial aspects of the CCA and ICA.

Lastly, stenosis was defined as the ratio of luminal to outer wall cross-sectional areas at locations where lumen was compromised as demonstrated in Figure 1. This type of measurement was preferred over the NASCET method as it allowed capturing small stenoses, i.e. when the lumen size was larger than the lumen of the distal ICA.

### Prediction of stenosis at follow-up

Our previous study has demonstrated differences in geometric remodeling of healthy and diseased carotid arteries with age^3^. Normal aging in the absence of ACD was associated with increase in carotid bifurcation angle and tortuosity, whereby carotids with ACD had straighter shapes and narrow bifurcation angles^3^. Since young healthy carotid arteries also typically have straight shapes and narrow bifurcation angles, all geometric features need to be considered in the context of age. To accomplish this, equations describing increases in bifurcation angle, CCA tortuosity, and bulb cross-sectional area with age in normal subjects without ACD were used to calculate “idealized” geometric parameters for healthy carotid arteries using the following relations^3^:

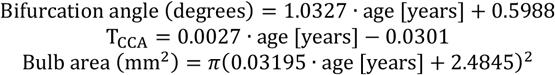

These “idealized” values were then compared against the actual bifurcation angle, tortuosity, and area measured using baseline CTA, and differences between the measured and calculated geometries were used in a multiple linear regression analysis along with clinical risk factors to predict stenosis severity at follow-up.

Age, gender, BMI, HTN, DM, dyslipidemia, CAD, tobacco use, and morphometric deviations in bifurcation angle, CCA tortuosity, and bulb area at baseline were used as independent variables. Analysis was performed with SPSS v22 (IBM, Armonk, NY) using stepwise linear regression to determine statistically significant predictors. A variable was entered into the model when the significance level of its F value was <0.05. Both unstandardized and standardized β coefficients were determined, and model quality was assessed with adjusted *R*^2^. Analysis was repeated excluding morphometric deviations and considering risk factors alone, and then excluding the risk factors and considering morphometric deviations alone. Finally, paired t-tests with p<0.05 considered statistically significant were used to determine if geometric features measured at follow-up were different from those measured at baseline.

## RESULTS

Average stenosis severity at baseline was 39±26% (range 0-89%). At follow-up stenosis increased to 44±28% (range 0-92%), and this increase was statistically significant (p=0.03) (Table 1) although no change over the follow-up was observed in 47% of arteries. Stenosis of >50% was present in 13 arteries at baseline, and in 15 at follow-up. Maximum increase in stenosis severity was 37% observed over 11 years (Figure 1B), while average increase was 5±12% observed over 4.7 years. Two representative cases demonstrating increases in stenosis severity over 4 and 11 years are presented in Figure 1 panels A and B respectively.

**Table 1.** Measured and calculated geometric features of the carotid arteries at baseline and follow-up. Calculated values were obtained for “idealized” healthy carotids using age as an independent variable. Differences between measured and calculated values at baseline were used in a multiple linear regression model to predict stenosis severity at follow-up.

|  | Measured |  |  | Calculated for “idealized” healthy carotid artery |  | Difference between measured and calculated at baseline |
| --- | --- | --- | --- | --- | --- | --- |
|  | Baseline | Follow-up | p | Baseline | Follow-up |  |
| Stenosis severity, % | 39±26 | 44±28 | 0.03 | | | |
| Bifurcation angle, $^\circ$ | 31±11 | 35±14 | 0.01 | 61±17 | 66±18 | 30±17 |
| T<sub>CCA</sub> | 0.08±0.06 | 0.10±0.07 | 0.01 | 0.13±0.04 | 0.14±0.05 | 0.05±0.05 |
| Bulb area, mm<sup>2</sup> | 61±20 | 62±19 | 0.60 | 61±14 | 65±15 | 1±26 |

Carotid artery bifurcation angle and tortuosity were larger at follow-up compared with baseline (31±11° vs 35±14° and 0.078±0.058 vs 0.099±0.072, respectively [both p=0.01]), but the largest cross-sectional area of the carotid bulb did not change (61±20 mm^2^ vs 62±19mm^2^ [p=0.60]). Calculated “idealized” bifurcation angles, CCA tortuosity, and bulb cross-sectional area for healthy carotid arteries were 61±17°, 0.13±0.04, and 61±14 mm^2^ at baseline, and 66±18°, 0.14±0.05, and 64±15 mm^2^ at follow-up, respectively. Baseline calculated values were on average 30±17°, 0.05±0.05, and 1±26 mm^2^ different than those measured using CTA (Table 1).

Older age and the presence of CAD and DM positively contributed to stenosis severity at follow-up, and explained 47% of variation in stenosis. Beta weights for the multiple linear regression model that used only risk factors as independent variables are provided in Figure 2A. Numeric values within the bars represent unstandardized weights, whereas bar lengths represent standardized weights that demonstrate the influence of each parameter on stenosis severity. When coupled with morphometric deviations in CCA tortuosity and carotid artery bifurcation angle, CAD, HTN, and Gender explained 67% of variation in stenosis severity at follow-up (Figure 2B). Female gender and risk factors had positive contributions to stenosis severity (note that Gender = 1 [male] results in smaller stenosis). Narrow bifurcation angles and large CCA tortuosity compared with “idealized” healthy values also contributed to more severe stenosis. Interestingly, age and DM lost their predictive values when analysis was performed including morphometric deviations, although age was used to calculate morphometric deviations in both angle and tortuosity. When using morphometric deviations alone, 23% of variability in stenosis severity at follow-up was explained by the difference in carotid artery bifurcation angle.

**Figure 2.**
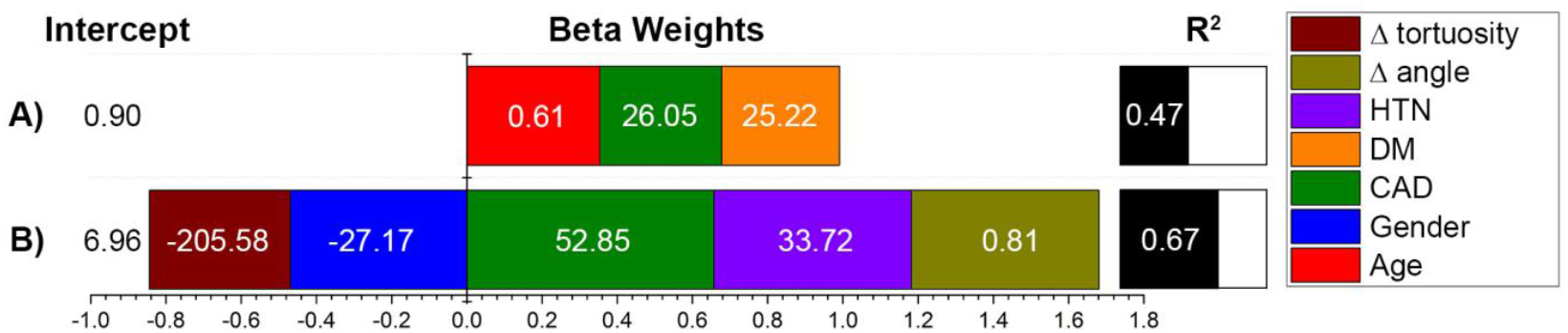
Stenosis severity at follow-up (%) can be predicted by patient demographics, risk factors, and differences in measured and calculated “idealized” geometric features at baseline. Unstandardized beta weights for the multiple linear regression models are provided with numeric values within the bars, whereas the length of the colored bars is made using standardized weights and represents the influence of each parameter on stenosis at follow-up. *R*^2^ demonstrates the quality of the model, i.e. how much variability in stenosis severity at follow-up is described by demographics, risk factors, and morphometry. Graph (A) represents prediction using only demographics and risk factors, while graph (B) also considers the influence of deviations in morphometry. Here tortuosity is dimensionless, angle is measured in degrees, HTN, DM, CAD are binary [1=yes, 0=no], gender is binary [0=female, 1=male], age is measured in years. For example, expected stenosis severity for a 50-year-old male with HTN and a bifurcation angle of 20° is 6.96 – 27.17·1[male] + 33.72·1[HTN] + 0.81·32[Δ angle] = 39%. Note that a bifurcation angle for a healthy 50-year-old patient is 1.0327·50[years] + 0.5988 = 52°, which is 32° larger than measured in our hypothetical subject.

## DISCUSSION

Geometry of the carotid artery undergoes significant changes with age^3,5–7^, and some of these changes are thought to be attributed to the characteristics of elastic fibers in the carotid artery wall^3,8,9^. Having formed during perinatal period^10^, elastic fibers stretch as vessel grows in length, resulting in significant tension in maturity. This tension, known as longitudinal pre-stretch, has a fundamental role in arterial growth and adaptation^8^, profoundly effecting changes in arterial shape and size with age^4^. Young healthy arteries are significantly pre-stretched longitudinally, which results in a straight geometry and narrow bifurcation angles^3^. With age, elastic fibers degrade and fragment, releasing the pre-stretch and resulting in longer, more tortuous arterial geometry and wider bifurcations^3^. Furthermore, muscular ICA and elastic CCA demonstrate differences in geometric remodeling, which may be associated with differences in elastic fiber orientations^4^. In the muscular ICA elastic fibers are oriented in the longitudinal direction, while in the elastic CCA they are both longitudinal and circumferential^11^. Release of tension due to degradation and fragmentation of elastic fibers with age therefore results in wider CCAs and more tortuous ICAs and CCAs. These findings clearly demonstrate the importance of age in studies of carotid artery geometry^3,5^, which have led to the development of formulas describing normal changes in carotid artery bifurcation angle, tortuosity, and diameter with age^3^.

In older subjects, some of these geometric features have also been demonstrated to be a risk factor for ACD^12,13^. Presence of disease was associated with wide bulbs, narrow bifurcation angles, tortuous CCAs, and straight ICAs^3^. Interestingly, while some of these features, like narrow bifurcation angles and straight ICAs, are usually found in young healthy carotid arteries, their presence in older subjects was an indicator of ACD, again emphasizing the need to consider arterial geometry in the context of age. These findings stimulated this current analysis, whereby geometry of the carotid artery obtained by clinical imaging at baseline was compared to normally aging “idealized” healthy artery to help predict ACD progression. Our results demonstrate that narrow bifurcation angles and tortuous CCAs in senior subjects are associated with increased stenosis severity over time. These results may help identify carotid geometries that have started undergoing abnormal remodeling, either due to onset of atherosclerotic disease, or other pathologic factors.

In order to make predictions more accurate, geometric features were combined with traditional Framingham risk factors that are known to be associated with the onset and progression of ACD. Indeed, when geometric and risk factor characteristics were combined, they were able to explain 67% of deviation in stenosis severity at follow-up, compared with only 47% when using risk factors alone, and 23% when using geometry alone. These results demonstrate the importance of a synergistic consideration of biologic and geometric variables in understanding and potentially predicting ACD progression, even though a 5±12% increase in stenosis severity over the 4.7 year follow-up observed in the current study may not be clinically significant.

Another interesting finding that emerged from this synergistic consideration of geometric and biologic variables was the influence of gender on stenosis progression. Carotid artery stenosis is known to be more prevalent in men than in women^14^, but women appear to have higher perioperative risk and higher incidence of recurrent disease^15^. Our data demonstrate that gender does not contribute to stenosis progression when analysis accounts for age, but does not account for the geometric characteristics of the carotid artery; however, when these geometric features are considered, the interplay of geometry and gender appears to become statistically significant. While this finding requires further evaluation with larger sample sizes, it suggests that female carotid arteries may progress to higher grades of stenosis faster than those of males if both arteries have similar geometry.

While prediction of stenosis severity at follow-up using morphometric features and traditional risk factors may hold promise in helping better identification of patients at higher risk for ACD, these results should be considered primarily as an illustration of the method, not as a definitive tool for measuring stenosis progression because we cannot exclude a type I error due to a small sample size. Larger analysis of patients with small carotid stenosis can be very instrumental in refining and validating the presented model, but these patients typically do not undergo clinical imaging, much less repeated imaging evaluations. Therefore, the pool of such patients at a single institution is quite small, which calls for coordinated efforts in image analysis. The second limitation is related to a relatively short average 5-year follow-up period used in this study. Due to generally slow nature of the carotid disease, longer time periods between the scans can greatly improve prediction accuracy. On the other hand, ACD can also progress non-linearly and rapidly due to a variety of factors including unstable plaques, intra-plaque dissection, inflammation, or thrombosis, which may significantly complicate stenosis prediction. This also calls for larger sample sizes that would allow expanding the range of considered factors to include structural plaque characteristics. While these limitations are being addressed, presented analysis can be instrumental in predicting uncomplicated ACD progression in small-grade stable carotid lesions, which may eventually lead to better predictive modeling and personalized treatment recommendations for patients with early stages of ACD.

## Data Availability

All data produced in the present study are available upon reasonable request to the authors

## REFERENCES

1. Benjamin EJ, Blaha MJ, Chiuve SE, et al. Heart Disease and Stroke Statistics—2017 Update: A Report From the American Heart Association.; 2017. doi:10.1161/CIR.0000000000000485.

2. Malek AM, Alper SL, Izumo S. Hemodynamics Shear Stress and Its Role in Atherosclerosis. JAMA 1999;282:21:2035-2042.

3. Kamenskiy A V., Pipinos II, Carson JS, MacTaggart JN, Baxter BT. Age and disease-related geometric and structural remodeling of the carotid artery. J. Vasc. Surg. 2014;In Press. doi:10.1016/j.jvs.2014.10.041.

4. Kamenskiy A, Miserlis D, Adamson P, et al. Patient demographics and cardiovascular risk factors differentially influence geometric remodeling of the aorta compared with the peripheral arteries. Surgery 2015;158([Epub ahead of print]):PubMed PMID 26096560; NIHMSID: NIHMS697336. doi:10.1016/j.surg.2015.05.013.

5. Thomas JB, Antiga L, Che SL, et al. Variation in the carotid bifurcation geometry of young versus older adults: implications for geometric risk of atherosclerosis. Stroke 2005;36(11):2450–2456.

6. Polak JF, Kronmal RA, Tell GS, et al. Compensatory increase in common carotid artery diameter. Relation to blood pressure and artery intima-media thickness in older adults. Cardiovascular Health Study. Stroke 1996;27(11):2012–2015.

7. Steinke W, Els T, Hennerici M. Compensatory carotid artery dilatation in early atherosclerosis. Circulation 1994;89(6):2578–2581.

8. Humphrey JD, Eberth JF, Dye WW, Gleason RL. Fundamental role of axial stress in compensatory adaptations by arteries. J. Biomech. 2009;42(1):1–8. doi:10.1016/j.jbiomech.2008.11.011.Fundamental.

9. Jackson ZS, Dajnowiec D, Gotlieb AI, Langille BL. Partial off-loading of longitudinal tension induces arterial tortuosity. Arterioscler. Thromb. Vasc. Biol. 2005;25(5):957–62. doi:10.1161/01.ATV.0000161277.46464.11.

10. Mithieux SM, Weiss AS. Elastin. Adv. Protein Chem. 2005;70(null):437–61. doi:10.1016/S0065-3233(05)70013-9.

11. Kamenskiy AVA, Dzenis YYA, Kazmi SAJSAJ, et al. Biaxial Mechanical Properties of the Human Thoracic and Abdominal Aorta, Common Carotid, Subclavian, Renal and Common Iliac Arteries. Biomech. Model. Mechanobiol. 2014;13(6):1341–59. doi:10.1007/s10237-014-0576-6.

12. Phan TG, Beare RJ, Jolley D, et al. Carotid artery anatomy and geometry as risk factors for carotid atherosclerotic disease. Stroke. 2012;43(6):1596–601. doi:10.1161/STROKEAHA.111.645499.

13. Bijari PB, Wasserman B a, Steinman D a. Carotid bifurcation geometry is an independent predictor of early wall thickening at the carotid bulb. Stroke. 2014;45(2):473–8. doi:10.1161/STROKEAHA.113.003454.

14. Stoberock K, Debus ES, Atlihan G, et al. Gender differences in patients with carotid stenosis. Vasa. 2016;45(1):11–6. doi:10.1024/0301-1526/a000490.

15. Kuy S, Seabrook GR, Rossi PJ, Lewis BD, Dua A, Brown KR. Management of Carotid Stenosis in Women. JAMA Surg. 2013;148(8):788. doi:10.1001/jamasurg.2013.342.

